# Development, evaluation, and implementation of a tablet device-based depression screening and management tool for rural women’s self-help groups

**DOI:** 10.1101/2024.11.06.24316834

**Authors:** Bharat Kalidindi, Abijeet Waghmare, Krishnamachari Srinivasan, Johnson Pradeep, Ramakrishna B Goud, Amritha Bhat, Dhinagaran Devadass, Tony D S Raj

## Abstract

Depression is a critical public health issue among women in rural India, with diagnosis and treatment rates being very low. The paper summarizes the development of MITHRA, a user-centred mobile app for depression screening and treatment among women in self-help groups (SHGs) of rural India. The predevelopment phase involved situation analysis and forming participatory design groups of prospective users. The app development phase used an Agile approach for flexibility and rapid adaptation. The post-development phase involved user acceptance testing, training on app usage, data synchronization, accuracy testing and usability evaluation. The study found that only 25% of the women in the participatory design group were digitally literate, despite 63% smartphone ownership. On assessing the cellular connectivity, the average download speed across the eight study sites was 9.81 Mbps, and the average upload speed was 4.6 Mbps. The user tests showed improvement in the task success rate between user testing session 1 and session 2 for all the tested tasks. The app successfully passed user acceptance testing (UAT) and data accuracy testing, demonstrating its readiness for deployment. Heuristic evaluation identified 52 violations with a mean score of 2.3, indicating minor usability issues to be addressed. This description of app development underscores the potential of mHealth apps in addressing mental health issues in underrepresented populations.

**What is already known on this topic:** Depression bears a high burden in rural India, particularly among women. Despite high prevalence, diagnosis and treatment rates remain low due to limited resources and stigma. Additionally, lack of awareness contributes to the problem, as many individuals in rural communities may not recognise depression as a treatable condition. Moreover, the scarcity of mental health specialists exacerbates the situation, especially in underserved rural areas.

**What this study adds:** This study underscores the importance of a human-centred app development processes, utilizing a participatory design group approach that places users at the core of every development phase. This study uniquely involved no-to-low literate women and poor digital literacy from rural India as the primary users. By integrating these users before, during, and after the app development phase, the study ensured that the app was intuitive and accessible, specifically catering to the needs of this demographic. The app’s design and functionality were directly influenced by the users, enhancing its effectiveness during mental health interventions. This approach improved usability, demonstrating the profound impact of inclusive and user-focused development practices.

**How this study might affect research, practice or policy:** The results from this study reveal that incorporating human-centred design through a participatory approach—where a group of women from the same rural community actively contributed to an app development—leads to the development of an app that is well accepted by the target population. These findings represent a significant advancement in healthcare technology, emphasizing the importance of user engagement and cultural relevance in creating effective solutions. As researchers, policymakers, and practitioners consider these results, they may reshape methodologies, policies, and clinical practices to prioritize user-centric approaches and enhance overall healthcare outcomes. This study also highlights a potential alternative, accessible treatment option for rural communities, partially addressing the mental health workforce shortage.

## Introduction

Depression is a prevalent mental health issue in India, with 20% of women affected with considerable barriers to access treatment. [1–4] This is due to several factors, including stigma, dearth of trained mental health practitioners, and cultural barriers associated with the subordination of women, low autonomy, and suppressed decision-making authority.[5,6] Early intervention for mental health can minimize hospital admissions, shorten hospital stays, improve acceptance/adherence, improve functioning and overall quality of life, and result in cost savings for healthcare providers.[7–10] Therefore, overcoming barriers to accessing mental health care among rural women is vital.

Mental health (mHealth) apps can reduce the burden on mental health services, improve patient well-being, and increase access for underrepresented populations, especially those with mild to moderate depression and anxiety. [11–13] These apps can identify mental health disorders early and support self-management, making them convenient, affordable, and applicable in various settings, including rural areas. [14,15] mHealth apps are scalable, empowering, and allow individuals to manage their mental health without the restrictions of traditional services. [16] Previous studies have shown that a stepped-care approach using mHealth can reduce the burden on healthcare systems by providing appropriate treatment to a wider population. [17,18] However, user engagement is low, with drop-out rates as high as 47% due to usability and network issues. [19] Despite mobile ownership in India being approximately 87%, usage of text messaging is only 14% [20], indicating that mobile based interventions may not be accessible to all. A collective tablet-based intervention for depression screening and management could help increase participation, adherence and support. [21–23]

A user-centred tablet-based app was developed using participatory design approach. [23] MITHRA (Multiuser Interactive Health Response Application) was developed to screen and track depression and deliver components of the evidence-based therapy, the Healthy Activity Program (HAP), for people with mild to moderate depression. [24] This paper describes the design and development process of the MITHRA app. This app was designed and developed for the rural women who are part of a homogenous self-help groups (SHGs) which are formed to empower them, socially and financially and are supported by local non-government organization (NGOs).

## Methods

### Development Phases

The MITHRA app aimed to enable women to use it offline at their convenience in the SHGs. The app development involved three phases: predevelopment phase, app and content development phase, and post-development phase. The activities during these phases are depicted in the Figure 01.

**Figure 01:**
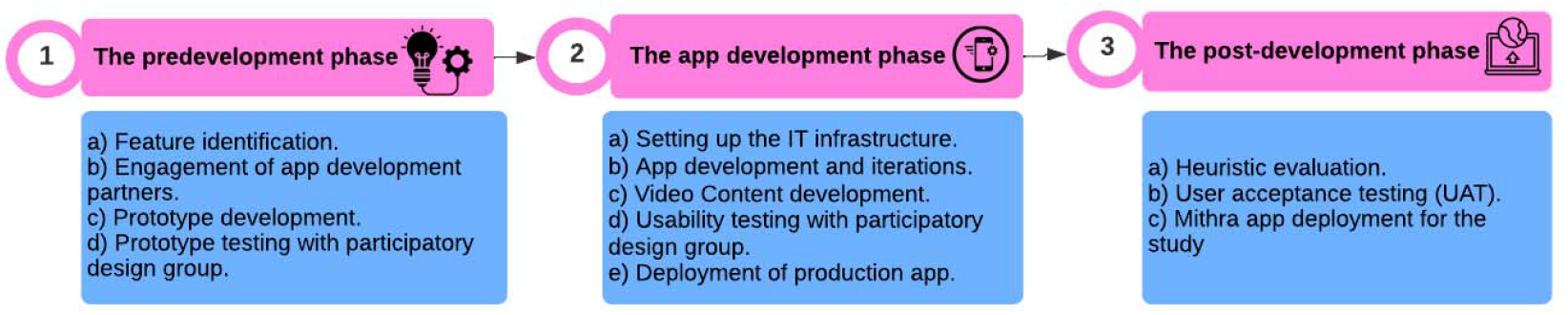
Phases and Activities.

### Predevelopment phase

#### Situation analysis at the study sites

Three SHGs were visited to understand the local context of the targeted user groups. SHG coordinators were interviewed to understand their workflows and get familiar with the participating members of the SHG. Two focus group discussions (FGDs) were conducted to understand the demographics, literacy levels, and current mobile app usage (Manuscript submitted for publication) of SHG members. Considering the poor to moderate network connectivity in rural areas, internet speed tests were conducted to understand possibilities of app usage and content streaming.

To protect the Tablet device, a strong protective case, and a portable power backup was provided for each SHG to extend use. Location tracking was enabled on the device and nonessential functions were disabled to promote focussed use. The MITHRA app has four user roles: participant, researcher, coordinator, and administrator. Participants undergo mental health screening and intervention through the app. Researchers conduct participant assessments blinded to study conditions. Coordinators register participants and oversee intervention status. Administrators manage study progress, create users, and manage the database. The features identified for these roles are shown in the Figure 02.

**Figure 02:**
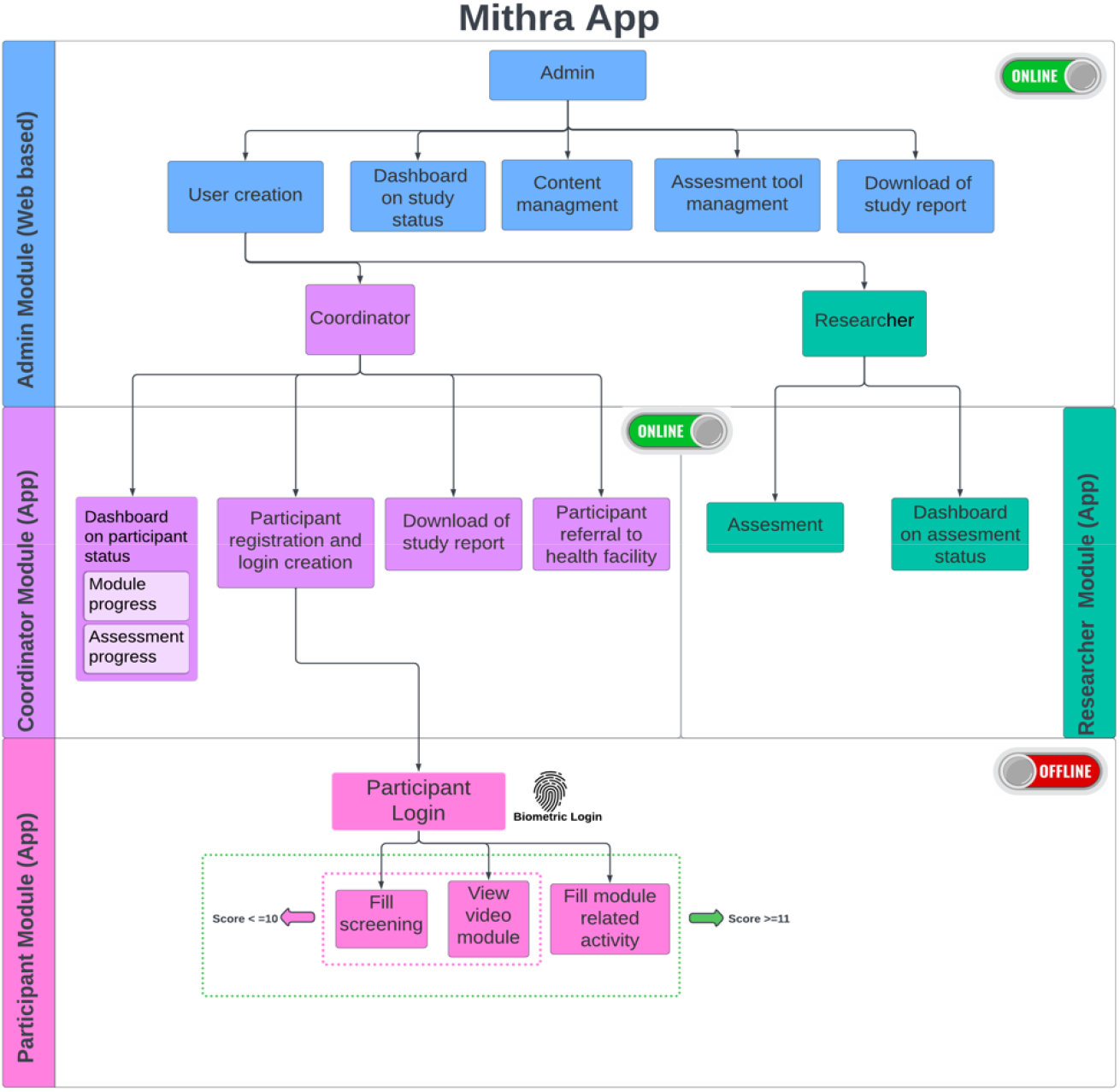
MITHRA app users and identified features required for the study.

#### Forming a participatory design group

Prior to starting the app development process, we conducted Focus Group Discussions (FGDs) with women from SHGs to understand the barriers and facilitators to community-based depression screening and treatment. A group of women from these FGDs were randomly selected to form a participatory design group (PDG). They provided inputs on app design, usability, and content appropriateness through basic app screen wireframes and video modules that were developed during this phase. They also discussed and finalised possible names for the animated characters, voice-over language, and accent in the educational videos.

### App and content development phase

#### MITHRA app development process

The MITHRA app was developed using Agile process, an iterative design method used for app development [25]. The app development process started with requirement analysis from the PDG, and based on this, app design wireframes were developed. App coding was initiated once the workflows were finalized. The study and app development team prioritised the required features for each iterative phase (sprint). Each sprint output was tested by the study team, suggested improvements were incorporated, and the errors (bugs) were identified and resolved.

#### Software technology and hardware

The MITHRA tablet app runs on Android OS 12 and uses a local SQLite database to store and retrieve data, allowing it to function offline. When internet is available, the app syncs and exchanges data automatically with the server, to stay up to date. The Apache web Server hosts the Flask framework with JINJA templates for the front-end display and Python for the backend logic[26]. Additionally, the Frappe framework running on Nginx Web Server uses MariaDB database to manage the data. Thereby, the MITHRA app architecture is optimized to provide offline and online usage. The MITHRA app architecture is shown in the Fig. 03.

**Figure 03:**
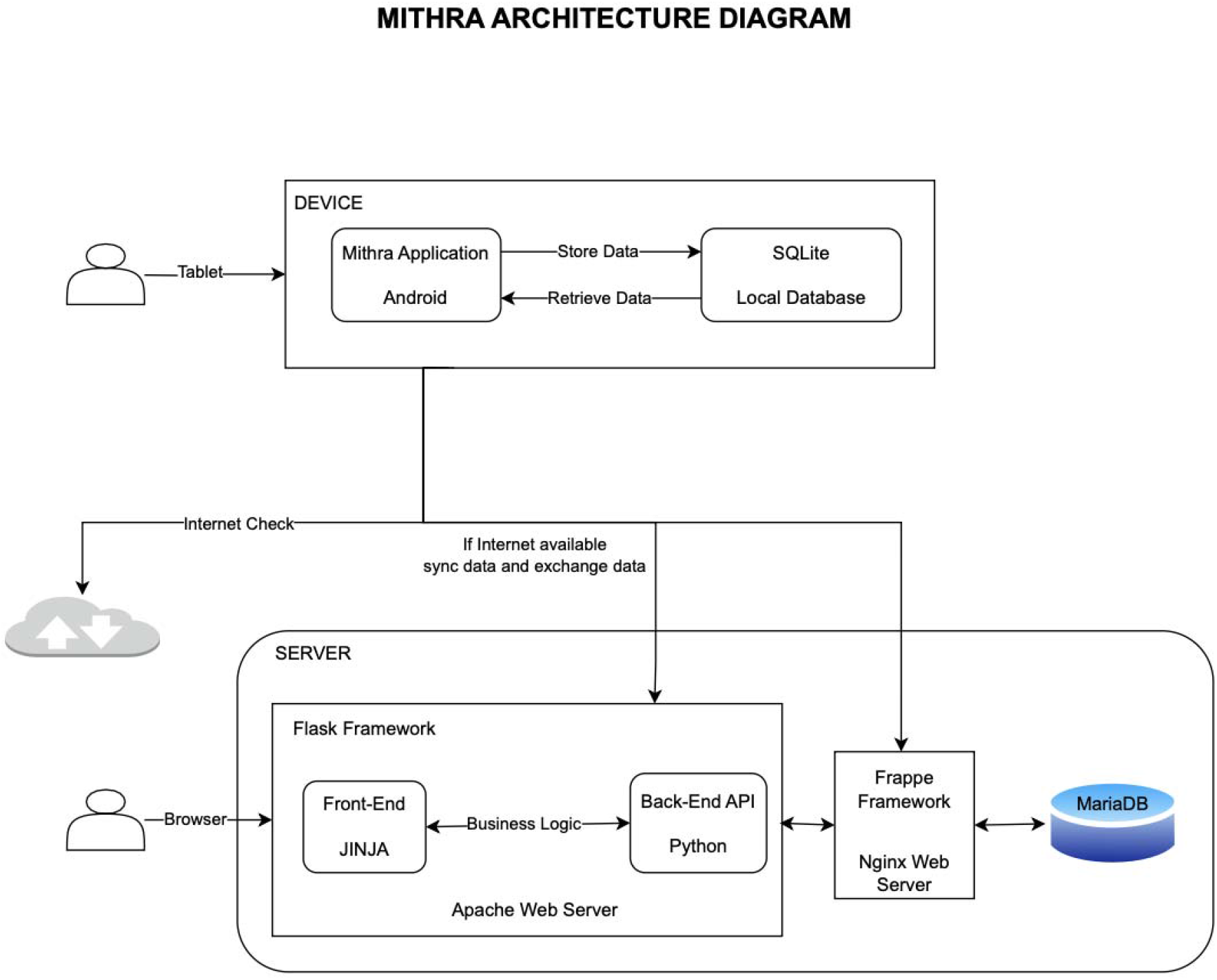
MITHRA app architecture diagram.

The Frappe Framework was chosen for application development due to its comprehensive, full-stack nature and preference for configuration over code. It offers built-in capabilities like authentication, database management, form builder, user roles, and permissions modules. Its ability to handle complex applications with a small team of developers was particularly beneficial for a mental health app like MITHRA making it ideal for customization and scalability.

The tablet hosting the app was a Samsung S6 Lite 10-inch tab with 4 GB RAM and 64GB ROM. The tablet had enhanced features, robust security updates, and a larger userbase. The 10-inch screen was selected as it provided a balance between portability and usability, enhancing user interaction. The Snapdragon processors enabled high performance and efficiency, ensuring smooth app operation and compatibility testing. The 4 GB RAM provided smooth functioning of resource-intensive tasks and a better user experience. The 64 GB storage was chosen as it provided ample space for the application’s data, resources, and updates. The inclusion of 5G, WiFi, and Bluetooth was necessary for high-speed data transfer and seamless internet connectivity. Additionally, a 20,000 mAh power backup extended the internal battery life (6000mAh) and an external fingerprint authenticator provided easy access.

#### Content Development

The application’s content was based on the HAP, an opensource brief psychotherapy developed by Sangath & The London School of Hygiene and Tropical Medicine. HAP aims to provide non-specialist Mental Health & Psychosocial Support providers with information to support patients with depression in primary care settings [27].

The HAP Modules were transformed into videos by creating storyboards for animation flows and colloquially translating dialogues into Kannada which were then recorded using a female voice. Animated characters were created using Macromedia software, with inputs from PDG participants on the animations and voice-overs. The intervention videos were programmed to appear sequentially based on the screening scores of users.

#### Data synchronization and monitoring

The MITHRA participant app is designed to function offline, thereby collecting and storing data on a tablet device when logged in as participant ensuring network issues don’t disrupt app usage. When the study coordinator logs in using the login credentials, app synchronizes with the online server when a network connection is available and participants’ data on app usage and progress from different study devices is synced back to the tablet device. The study coordinator can synchronize data, apply updates, and manage content on the tablet device. The first installation on each tablet requires coordinator’s login to load the application and media content and there on the participant can operate the app entirely offline. The app synchronization flow is shown in the Fig 04.

**Figure 04:**
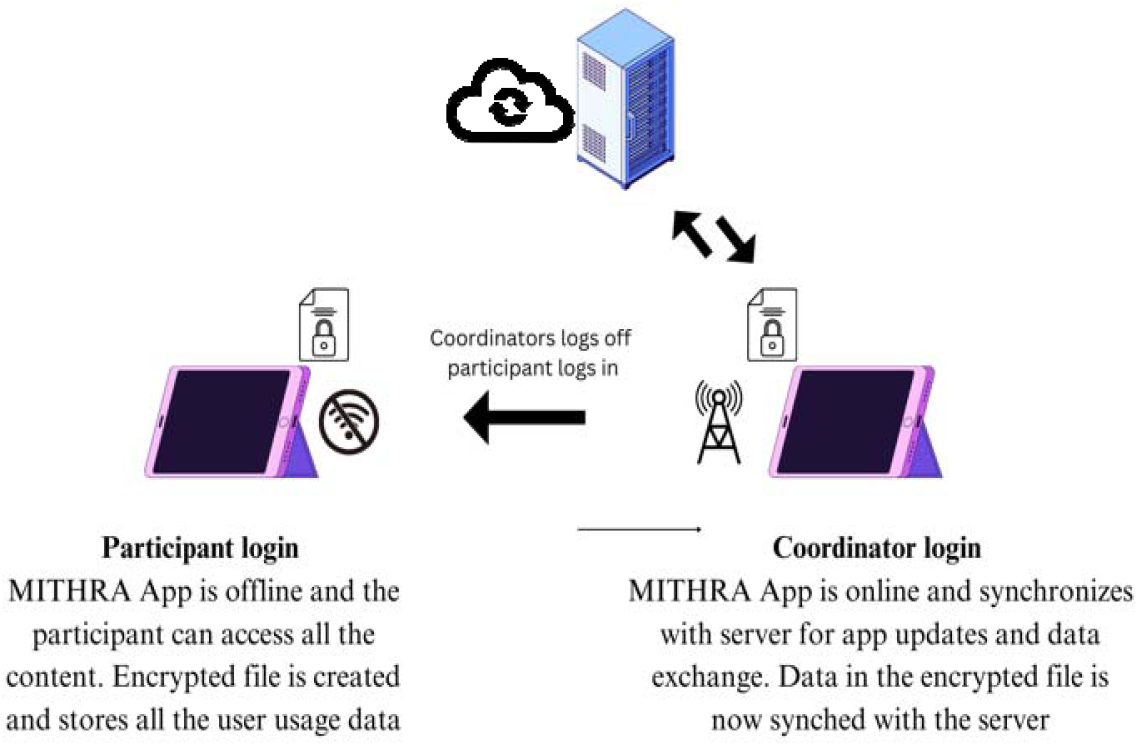
Mithra App data synchronization and monitoring process.

### Post development phase

#### System and Data accuracy testing

After development, extensive data accuracy testing was conducted on the MITHRA app using five synthetic datasets that simulated real world scenarios. Functional testing of the application and unit testing of all modules were performed and documented. Following deployment, periodic monitoring of data synchronization between the tablets and servers was maintained.

#### User Acceptance Testing (UAT)

The UAT was conducted to ensure the full functionality and usability of the MITHRA app before its deployment. Key users were identified from PDGs, and a UAT plan was developed with testing objectives, scenarios, and success criteria for each user group. Realistic case scenarios were piloted during training sessions to familiarize users with the application’s interface and functionality. Feedback was collected from users regarding the application’s ease of use, performance, and overall usefulness which was analyzed to identify trends and issues. The development team addressed these issues, and the updated application was retested to ensure successful resolution. Once all issues were resolved and the application met acceptance criteria, the UAT was signed-off, confirming its readiness for deployment.

#### App usage and device management training

Members of the SHG were trained on basic tablet device use, biometric authentication, data privacy, login and logout processes, app layout, media player controls, and icons and action buttons. Training took place in November 2022, with individual technical assistance sessions for those needing extra training. Members who demonstrated high comfort and confidence were identified as champions and they demonstrated app usage to other members. The training sessions lasted around 60 minutes at each study site. Each champion was responsible for storing, charging, and handing the device to any SHG member who wanted to use the MITHRA app. They were provided with a device protection case, charging equipment, and a power backup for safe and uninterrupted device usage.

#### Evaluation of usability heuristics

The MITHRA app was evaluated using Nielsen’s usability heuristics [28] which involve identifying heuristic violations and their severity by evaluators who understand user needs, human-computer interaction, interface design, cognitive and perceptual psychology and information architecture. Four evaluators participated in the study, each receiving orientation on the heuristic evaluation, steps to carry out the evaluation, and viewing 30 minutes of video content from the Nielsen-Norman Group website. They accessed the app on an Android-based tablet device, took screenshots of violations, and noted them in an Excel file for further assessment and necessary changes.

#### Data security, privacy, and confidentiality

All the users were given login credentials and biometric authentication to maintain data privacy. Personal headphones were used to use app modules. Only the study administrator had access to downloaded reports, including demographics, assessment responses, and app usage data. The administrator maintained a log on data usage and sharing.

## Results

### Pre-Development Phase

#### Findings of situational analysis

Participatory design group (PDG) was formed by randomly selecting 15 women from various villages that were a part of study geography. The participants demographics, smartphone ownership and digital literacy details are shown in the table no. 01.

**Table no. 01.** Details of PDG participants (n=16)

| Details | Frequency (%) |
| --- | --- |
| **Age** |  |
| 18-30 Years | 5 (31%) |
| 31-45 Years | 7 (44%) |
| 46 Years and above | 4 (25%) |
| **Education** |  |
| No formal education | 4 (25%) |
| Primary education | 7 (44%) |
| Secondary education and above | 5 (31%) |
| **Occupation** |  |
| Housewife | 5 (31%) |
| Agriculture | 6 (38%) |
| Others | 5 (31%) |
| **Digital literacy and smartphone ownership** |  |
| Digital Literacy (Understanding basic usability of smartphones) | 4 (25%) |
| Ownership of Smartphones | 10 (63%) |

Internet speed test was conducted on every study site using Speedtest by Ookla mobile app and the test results were noted. The average download speed across the eight study sites was found to be 9.81 Mbps, with average upload speed at 4.6 Mbps. The test results were as on March 2022 and these findings were crucial for understanding the feasibility of implementing the MITHRA app in the rural areas.

### Development Phase

#### Participatory Design Group Findings

Three PDG sessions were conducted, the initial PDG was to understand the user’s perspective, while the other two PDGs were for conducting the user tests using a structured tool with predefined scenarios. The scenario vide task success rate in both the PDG sessions is shown the Table 02.

**Table 02:** Task success rate during the two PDGs.

|  | Task success rate<br>(No. of participants who successfully completed the assigned task)<br>n=16 |  |
| --- | --- | --- |
| Scenario | Session 1 Task success rate | Session 2 task success rate |
| Login & Logout | 13 (81%) | 15 (93%) |
| Watch the MITHRA intervention video | 15 (93%) | 16 (100%) |
| Navigate through past and pending MITHRA intervention videos | 10 (62%) | 12 (75%) |
| Fill the activities done over past 1 week and activities willing to do over next 1 week | 3 (18%) | 7 (43%) |

### Post Development phase

#### UAT

Post development, the UAT for the MITHRA app was done and several test scenarios were executed to ensure the app’s functionality and usability. The brief description of test results are shown in table 03.

**Table 03:** User acceptance test scenarios and results.

| **Test Case ID** | **Test Scenario** | **Test Steps** | **Expected Result** | **Actual Results Status** |
| --- | --- | --- | --- | --- |
| TC01 | App Installation | Install the app on a smartphone | App is installed successfully | Pass |
| TC02 | User Registration | Register a new user with valid details | User is registered successfully | Pass |
| TC03 | User Login | Login with valid credentials | User is logged in successfully | Pass |
| TC04 | Access Intervention | Access video modules section in the app | Resources are displayed correctly | Pass |
| TC05 | Use Interactive Features | Use interactive features like self-assessment tools and activity tracker | Features work as expected | Pass |
| TC06 | Offline Accessibility | Access app features in offline mode | Essential features are accessible offline | Pass |
| TC07 | Multilingual Support | Change app language to a regional language | App interface changes to the selected language | Pass |
| TC08 | Uninstallation | Uninstall the app from the smartphone | App is uninstalled successfully | Pass |

#### Data accuracy testing

The data accuracy testing for the MITHRA app was conducted using a simulation dataset of five users that consisted of all the 314 variables that the app intended to collect during live deployment. The dataset covered all possible scenarios that could occur during the intervention. The data accuracy testing results showed that all the data points that the app collected were found to be 100% accurate and consistent. The MITHRA app’s data accuracy was confirmed by comparing input values with expected output, indicating its reliability in collecting and reporting user data.

#### Heuristic Evaluation

There were 52 heuristic violations identified on the MITHRA app that was used by members, researchers and coordinators in the tablet devices. In the Figure. 05, the vertical axis represents the Nielsen heuristics that were violated across the app and the horizontal axis represents the counts of violations.

**Figure 05:**
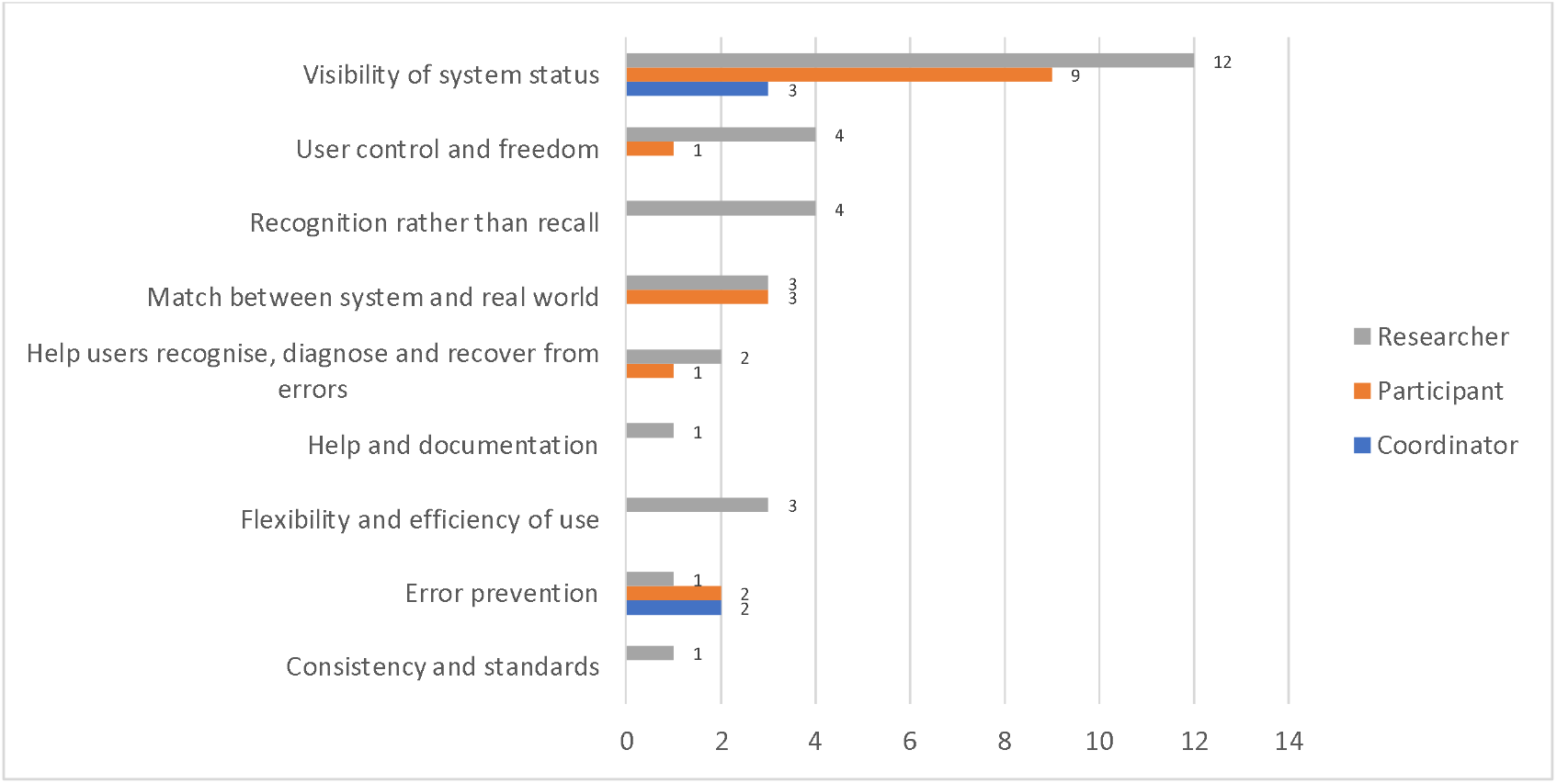
Count of heuristic violations by the MITHRA app.

The severity of the 46 heuristic violations ranged from 2.0 to 3.0 with a mean of 2.3 which refers to app having minor usability problems and these problems could be fixed on lower priority. Examples of heuristic violations by the MITHRA app is shown in the table no. 04.

**Table 04:** Sample description of heuristic violations by the MITHRA app.

| Usability heuristic | Mean of severity score (SD) | Sample description of violations in the Mithra app | Mithra app user type |
| --- | --- | --- | --- |
| Consistency and standards | 2 (0) | Survey completion status is inconsistently shown in the quick filters | Coordinator |
| Error prevention | 2.6 (0.5) | No error message when the researcher misses few survey questions. | Researcher |
| Flexibility and efficiency of use | 2.3 (0.5) | After finishing a survey for "Participant", the screen returns to the unfiltered list of participants, and I have to scroll down and find "Participant" | Researcher |
|  |  | again. |  |
| Help and documentation | 3.0 (0) | Details of the members whose survey was missed during the window period are not visible. Some help or guide feature would be helpful to know where to get this information from. | Researcher |
| Help users recognize, diagnose, and recover from errors | 2.7 (0.5) | Don't know how to go to the correct activities section. When I open activities after watching 1 video, it only shows what activities I did last week, does not show me activities I should do next week. Am I supposed to watch all the videos, including what is pending? Will take me too long to do that now. Not sure what to do. | Participant |
| Match between system and real world | 2.2 (0.4) | The language of certain questions in the surveys doesn't change during the app language change | Researcher |
| Recognition rather than recall | 2.3 (0.5) | Details of the members whose survey was missed during the window period is not visible | Researcher |
| System status | 2.5 (0.5) | If a member is opened from an Unfiltered list or "Pending" filtered list, then it's not clear which of the surveys are low priority. | Researcher |
| User control and freedom | 2.4 (0.5) | Login page does not allow the members to select the language of the screen | Participant |
| Visibility of system status | 2.2 (0.6) | Pressing Skip and Save appear to do the same thing. In both, the data was cleared from the screen and the same activity window was shown | Participant |

## Discussion

The study provides the rationale and methods of designing and developing a mobile mental health app for screening and delivering intervention for depression to the women in Self-Help Groups (SHGs) in the rural India who do not have access to mental health services. The development of a mental health app for rural Indian women using a user-centered design approach has highlighted the critical role of understanding the unique needs of underserved populations in creating effective digital health solutions.

While there is a growing body of research on mobile app interventions for mental health, studies specifically targeting rural women remain limited. This gap is particularly concerning given the unique challenges faced by women in these communities, such as limited access to healthcare, cultural stigma, and economic hardship. This study fills a significant gap in the literature, as there is limited research on mobile mental health interventions specifically tailored to rural Indian women.

Designing an app for women in self-help groups poised several challenges due to low digital literacy levels, limited privacy, and limited internet connectivity in rural areas.[29,30] The app needed an intuitive interface to reduce cognitive load for first-time users and to be lightweight and functional in low bandwidth and network scenarios. By directly engaging with the target users throughout the design and testing phases, the app was able to address specific barriers faced by rural women, including low literacy levels, cultural stigma surrounding mental health, and limited access to healthcare services. [29,30] User feedback from initial testing phases underscored the success of several key design elements. The app’s use of audio and visual aids, rather than text-heavy content, made it more accessible to women with low literacy levels. Furthermore, integrating localized, culturally relevant content resonated well with the users, making the mental health interventions more relatable and practical for daily use. These insights reinforce the importance of tailoring health technologies to the cultural and socio-economic realities of their intended audience. The collaborative process fostered a sense of ownership among the women participants, increasing their willingness to use the app and engage in self-care practices. This highlights the potential of participatory design in creating more inclusive digital mental health solutions for underserved population. The heuristic evaluation conducted on the application identified several minor usability issues. While these issues did not significantly impact the core functionality, they highlighted areas for improvement to enhance the user experience. These findings will guide refinements in the next iteration of the application, which will be developed as part of a larger, more comprehensive study.

The development team faced challenges with biometric authentication for women, requiring alternative, secure methods like facial recognition. Many women had faded fingerprints, reducing the effectiveness of fingerprint recognition technology. This raised concerns about inclusivity and appropriateness of such technology for this demographic. The experience underscored the need for understanding user context and physical conditions when designing digital health solutions. These challenges highlighted the context-specific, user-centric approach which were crucial in developing a mental health app for rural women in India, ensuring accessibility and usability.

In future iterations. MITHRA app can be integrated with wearable devices to track physical activity and sleep patterns, providing real-time data on users’ activity and sleep quality.

In conclusion, the development of the MITHRA app was a unique and iterative process that involved a participatory design approach, understanding the local context, incorporating user feedback, and conducting rigorous testing. The app has the potential to provide valuable mental health support to women in self-help groups and in other communities particularly in rural areas with limited network connectivity.

## Data Availability

All data produced in the present study are available upon reasonable request to the authors

## Acknowledgement

We extend our sincere gratitude to the Director and field coordinators of Jnana Jyothi, Anekal Bangalore, for their unwavering support in the implementation of this study across their practice areas. Special thanks to Mr. Vincy Prem and Mr. Tejesh S, our dedicated research associates, for their exceptional coordination of the study activities. We also wish to acknowledge Mr. Deepak Bhaskar and his team at ProbePlus for their invaluable contribution in developing the application for this study. We would like to acknowledge the authors team from Sangath and London School of Hygiene and Tropical Medicine who have developed ‘Healthy Activity Program’. Their collective efforts and collaboration have been instrumental in the success of this research.

## Ethical considerations

Regulatory and Institutional Review Board (IRB) approvals for the research study were obtained in advance from the respective institutional ethics committees (IEC approval number STUDY 00010415) and conducted in accordance with accepted national and international standards. The Indian Council of Medical Research’s Health Ministry Screening Committee also approved the main study.

